# Immunogenicity of heterologous prime/boost inactivated and mRNA COVID-19 vaccine

**DOI:** 10.1101/2021.11.20.21266644

**Authors:** Nasamon Wanlapakorn, Ritthideach Yorsaeng, Harit Phowatthanasathian, Nungruthai Suntronwong, Sitthichai Kanokudom, Natthinee Sudhinaraset, Yong Poovorawan

## Abstract

**Introduction:** In August 2021, Thailand imported the BNT162b2 mRNA COVID-19 vaccine. The prioritised group to receive the BNT162b2 vaccine were health professionals. The BNT162b2 vaccine scheduled for healthcare workers were two-dose regimen administered three weeks apart, the third dose booster in two-dose inactivated CoronaVac vaccine recipients or as a second dose in health professionals who had received the CoronaVac or adenoviral-vectored (ChAdOx1-S) vaccine as the first dose regardless of the interval between the first and second dose.

**Methods:** This study aims to evaluate the immunogenicity of the heterologous prime boost CoronaVac followed by BNT162b2 in health professionals.

**Results:** The CoronaVac/BNT162b2 vaccine recipients elicited higher neutralizing activity against the original Wuhan and all variants of concern than in the recipients of the two-dose CoronaVac.

**Conclusions:** The heterologous CoronaVac/BNT162b2 could be used as an alternative regimen in countries experiencing the vaccine shortages and in individuals experiencing the adverse events following CoronaVac.

## Introduction

In August 2021, Thailand imported the BNT162b2 mRNA COVID-19 vaccine. The prioritised group to receive the BNT162b2 vaccine were health professionals. The BNT162b2 vaccine scheduled for healthcare workers were two-dose regimen administered three weeks apart, the third dose booster in two-dose inactivated CoronaVac vaccine recipients or as a second dose in health professionals who had received the CoronaVac or adenoviral-vectored (ChAdOx1-S) vaccine as the first dose regardless of the interval between the first and second dose.

It is possible to mix and match vaccines in specific situations such as a vaccine shortage or adverse reactions following vaccine administration. The interval between the first and the second dose may vary according to the availability of a vaccine. This study aims to assess and provide preliminary data on the immunogenicity of heterologous prime/boost inactivated vaccine followed by the BNT162b2 mRNA vaccine at different intervals amongst Thai health professionals.

## Methods

We performed a cross-sectional study in which leftover sera samples from participants seeking antibody testing following vaccination at the Center of Excellence in Clinical Virology, Faculty of Medicine, Chulalongkorn University between September and October 2021 were further analysed. Only samples from participants who received heterologous prime/boost inactivated CoronaVac followed by BNT162b2 vaccine (hereafter referred to as CV/BNT162b2) were used. The study protocol was approved by the Research Ethics Committee of the Faculty of Medicine, Chulalongkorn University (IRB 870/64).

Two reference groups of Thai individuals vaccinated with two-dose CoronaVac (hereafter referred to as CV/CV) (n=170) and BNT162b2 regimen (hereafter referred to as BNT162b2/BNT162b2) (n=19) were included in the analysis.

Venous blood samples were collected between 21-35 days after the second dose vaccination and tested for SARS-CoV-2 spike receptor-binding domain (RBD) IgG by SARS-CoV-2 IgG II Quant assay (Abbott Diagnostics, Abbott Park, IL), SARS-CoV-2 spike RBD total immunoglobulin (Ig) by Elecsys SARS-CoV-2 S (Roche Diagnostics, Basel, Switzerland), and anti-spike protein 1 (S1) IgA by an enzyme-linked immunosorbent assay (ELISA) (Euroimmun, Lübeck, Germany).

The neutralizing activity was tested against the original Wuhan strain and variants of concern, B.1.1.7, B.1.617.2, and B.1.351, by an ELISA-based surrogate virus neutralization test (sVNT); cPass™ SARS-CoV-2 neutralizing antibody detection kit (GenScript Biotech, Piscataway, NJ). The methods were described in a previous study [1].

The differences in antibody responses between groups were calculated using the Mann-Whitney U test. A *p*-value <0.05 was considered statistically significant.

## Results

There were significantly more women in the CV/BNT162b2 group (Table 1) due to the fact that healthcare professionals enrolled in this study were mostly female nurses. Furthermore, the CV/BNT162b2 group was younger than the CV/CV group (χ^2^ test *p*-value <0.001). Unlike the homologous CV/CV and BNT162b2/BNT162b2 vaccination cohorts, there were variations in intervals between the first and second dose vaccinations among the heterologous CV/BNT162b2 vaccinees. We analysed the immunogenicity data of the heterologous CoronaVac/BNT162b2 vaccinees in two sets. The first set were those who received two vaccines 21-28 days apart, now termed the short-interval CV/BNT162b2 regimen, and the second set were those who received two vaccines for more than 7 weeks apart, now the termed long-interval CV/BNT162b2 regimen (Table 2).

**Table 1.** Characteristics of the participants in all vaccinated groups.

| Characteristics | CV/CV<br>(n = 170) | Short-interval<br>CV/BNT162b2<br>(n = 66) | Long-interval<br>CV/BNT162b2<br>(n = 10) | BNT162b2/BNT162b2<br>(n = 19) |
| --- | --- | --- | --- | --- |
| Sex, Female no. (%) | 89 (52.4%) | 63 (93.9%) | 8 (80.0%) | 12 (63.2%) |
| Age years, mean [SD] (min-max) | 42.3 [9.6]<br>(18.0-59.0) | 24.1 [6.2]<br>(20.0-49.0) | 31.1 [11.2]<br>(20.0-57.0) | 34.4 [11.9]<br>(14.0-54.0) |
| Interval between the 1<sup>st</sup> and 2<sup>nd</sup><br>dose (days) | 23.0<br>[21.0, 26.0] | 21.0<br>[21.0, 21.0] | 107.5<br>[77.75, 137.5] | 21.0<br>[21.0, 24.0] |
| Median [IQR] (min-max) | (21.0-28.0) | (21.0-26.0) | (50.0-156.0) | (21.0-31.0) |
| Interval between the last dose<br>and blood collection (days) | 29.0<br>[28.0, 31.0] | 29.0<br>[29.0-35.0] | 30.50<br>[29.0, 35.0] | 34.0<br>[31.0, 35.0] |
| Median [IQR] (min-max) | (27.0-49.0) | (21.0-35.0) | (29.0-35.0) | (31.0-35.0) |

**Table 2.** The geometric mean titer (GMT) of anti-receptor-binding domain (RBD) Ig and IgG, anti-spike protein 1 IgA and percentage inhibition against wild type and variants of SARS-CoV-2 in serum from individuals who received homologous CoronaVac (CV/CV), heterologous CoronaVac followed by BNT162b2 (CV/BNT162b2), and homologous BNT162b2 (BNT162b2/BNT162b2) vaccines.

| Results | CV/CV<br>(n = 170) | Short-interval CV/BNT162b2<br>(n = 66) |  |  | Long-interval<br>CV/BNT162b2<br>(n = 10) | BNT162b2/BNT162b2<br>(n = 19) |
| --- | --- | --- | --- | --- | --- | --- |
|  |  | Total (all age) | 18-29 years old<br>(n = 52) | 30-50 years old<br>(n = 14) |  |  |
| Anti-RBD Ig, U/mL | 97.9 | 1042 | 1051 | 1010 | 10485 | 1963 |
| GMT [95% CI] | [82.6, 116.1] | [828.6, 1311] | [815.8, 1355] | [550.9, 1852] | [7228, 15209] | [1378, 2798] |
| (range) | (0.40-1028) | (116.4-12865) | (116.4-8891) | (224.5-12865) | (4894-19966) | (314.2-4310) |
| Anti-RBD IgG, BAU/mL | 128.0 | 1475 | 1613 | 1058 | 2683 | 1651 |
| GMT [95% CI] | [113.7, 144.1] | [1267, 1716] | [1372, 1895] | [723.5, 1547] | [2075, 3469] | [1182, 2306] |
| (range) | (14.7-961.4) | (181.5-4303) | (181.5-4303) | (225.7-3911) | (1332-4827) | (279.4-5169) |
| anti-S1 IgA, OD/CO ratio | 0.88 | 6.55 | 7.25 | 4.49 | 9.00 | 5.62 |
| Median [IQR] (range) | [0.55, 1.79] | [4.87, 9.00] | [5.43, 9.00] | [3.05, 7.85] | [7.79, 9.00] | [4.51, 8.37] |
|  | (0.15-9.00) | (1.14-9.00) | (1.14-9.00) | (1.25-9.00) | (3.44-9.00) | (2.01-8.37) |
| sVNT (Wild-type), | 66.60 | 95.45 | 95.45 | 95.30 | 97.35 | 97.40 |
| % Inhibition | [48.86, 79.41] | [93.00, 96.63] | [93.60, 96.68] | [92.65, 96.78] | [96.55, 97.55] | [96.30, 97.70] |
| Median [IQR] (range) | (21.13-92.50) | (64.20-98.20) | (64.20-97.60) | (89.90-98.20) | (96.20-98.20) | (83.50-98.10) |
| sVNT (B.1.1.7), % Inhibition | 42.11 | 84.70 | 85.40 | 80.30 | 97.85 | 93.70 |
| Median [IQR] (range) | [28.97, 58.31]<br>(7.21-73.15) | [78.53, 88.60]<br>(45.60-97.80) | [78.90, 88.60]<br>(45.60-97.50) | [75.75, 86.70]<br>(65.40-97.80) | [97.20, 98.00]<br>(96.50-98.00) | [88.30, 95.10]<br>(54.90-96.30) |
| sVNT (B.1.351), % Inhibition | 34.86 | 77.25 | 76.80 | 77.25 | 95.40 | 88.60 |
| Median [IQR] (range) | [20.46, 47.33]<br>(4.15-72.43) | [69.28, 82.53]<br>(45.30-96.70) | [69.63, 82.58]<br>(45.30-95.80) | [64.98, 82.23]<br>(57.50-96.70) | [93.98, 96.25]<br>(89.30-97.00) | [80.30, 91.20]<br>(52.20-93.60) |
| sVNT (B.1.617.2), % Inhibition | 48.93 | 92.95 | 93.15 | 89.00 | 97.35 | 97.10 |
| Median [IQR] (range) | [36.12, 63.42]<br>(12.46-85.45) | [89.08, 94.95]<br>(67.70-98.00) | [90.90, 95.10]<br>(67.70-98.00) | [84.50, 91.83]<br>(83.40-97.30) | [96.55, 97.93]<br>(86.50-98.00) | [95.00, 97.60]<br>(68.70-98.10) |

Analysis of RBD-specific total Ig following a two-dose vaccination showed that regimens including BNT162b2 elicited higher responses compared to the homologous CoronaVac regimen (Figure 1A). Within the CV/BNT162b2 regimen, the long-interval regimen elicited higher RBD total Ig following the second dose vaccination compared to the short-interval regimen.

**Figure 1.**
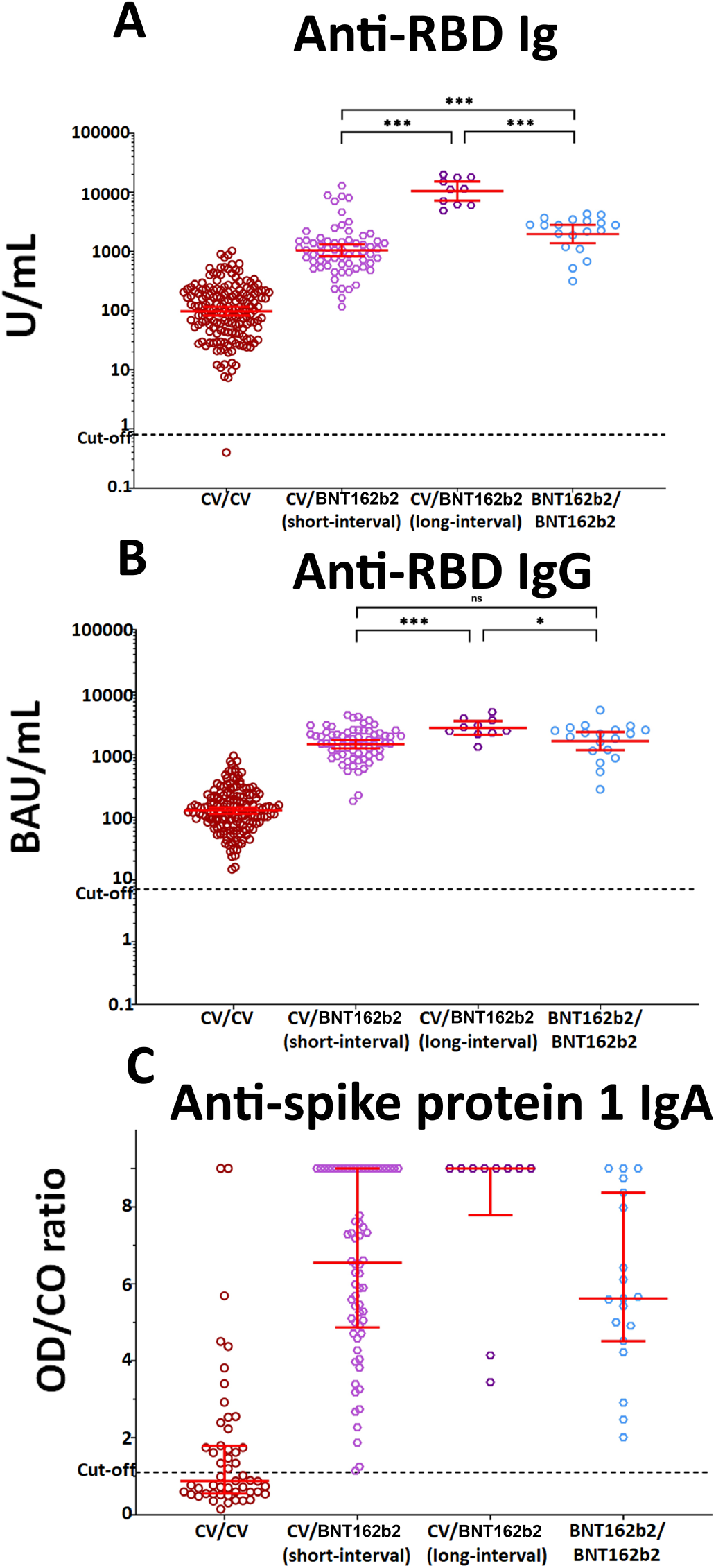
Binding antibody specific for SARS-CoV-2. A) Total immunoglobulin specific to the RBD (anti-RBD Ig) and B) Anti-RBD IgG and C) Anti-spike protein 1 IgA (anti-S1 IgA) at 21-35 days after completion of two-dose vaccination. Data points are the reciprocals of the individual. Lines indicate geometric means and bars indicate 95% confidence intervals for anti-RBD Ig and anti-RBD IgG, and median and interquartile ranges for anti-S1 IgA. Dotted lines denote the positive cut-off levels according to the manufacturer’s instructions. *** indicates *p* ≤ 0.001, ** indicates *p* ≤ 0.01. * indicates *p* ≤ 0.05, ns denotes no statistical significance.

When only considering anti-RBD IgG, all BNT162b2-incorporated regimens also showed higher anti-RBD IgG levels compared to the homologous CoronaVac regimen (Figure 1B). The long-interval CV/BNT162b2 regimen elicited higher anti-RBD IgG than the short-interval CV/BNT162b2 and homologous BNT162b2 regimens.

In addition, anti-S1 IgA was detected in only BNT162b2-incorporated vaccination schedules at similar levels, while no significant amount of anti-S1 IgA was detected in homologous inactivated CV/CV regimen (Figure 1C).

Comparison of neutralizing activities (presented as percent inhibition) showed that the long-interval CV/BNT162b2 regimen elicited higher neutralizing activities against the wild-type and all variants of concerns than the short-interval CV/BNT162b2 regimen (Figure 2A-D). The long-interval CV/BNT162b2 regimen also elicited higher neutralizing activities against the alpha and beta SARS-CoV-2 variants compared to the BNT162b2/BNT162b2 regimens (Figure 2B-C).

**Figure 2.**
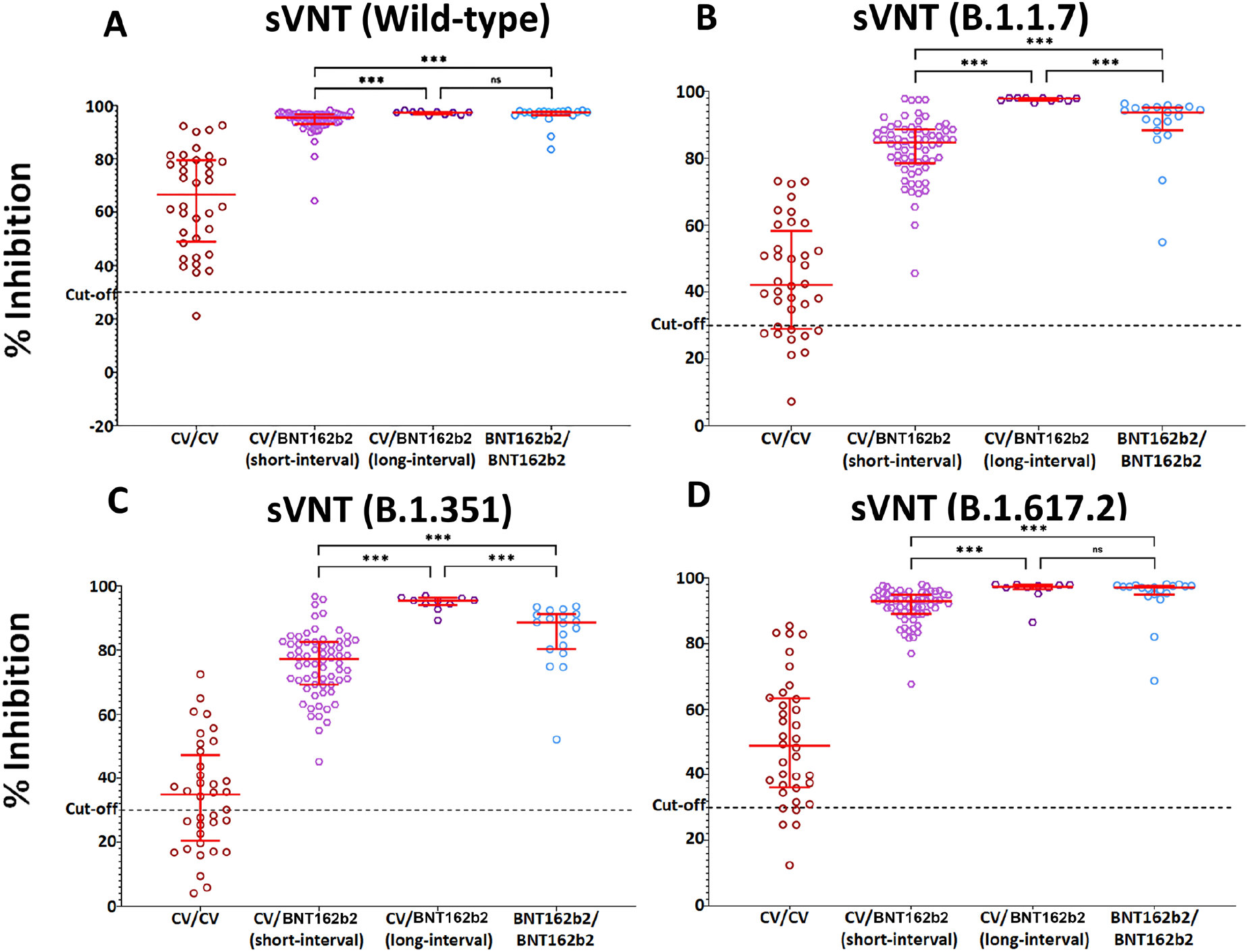
Serum neutralizing activities against wild-type SARS-CoV2 (D) and variants of concern (alpha (E), beta (F), and delta (G)) in vaccinated individuals at 21-35 days after completion of two-dose vaccination. The data points are the reciprocals of the individual. Lines indicate median and I-bars indicate interquartile ranges. Dotted lines denote the positive cut-off levels according to the manufacturer’s instructions. sVNT denotes surrogate virus neutralization test (sVNT). *** indicates *p* ≤ 0.001, ns denotes no statistical significance.

## Discussion

Our study found that both the short and long-interval heterologous regimens with CoronaVac followed by BNT162b2 induced higher SARS-CoV-2 RBD-specific antibody responses and neutralizing activities against wild type and variants of concern than that of the licensed two-dose CoronaVac vaccine with proven 65–83% efficacy against symptomatic COVID-19 [2, 3]. In addition, the SARS-CoV-2 RBD-specific total Ig and IgG responses elicited by CoronaVac/BNT162b2 were comparable to those elicited by homologous two-dose BNT162b2 vaccine. Although the antibody levels alone do not directly correlate with certain levels of protection, a previous study showed that higher anti-spike IgG, anti-RBD IgG, and neutralizing antibody titres are all associated with a lower risk of symptomatic COVID-19 [4].

Comparison between the short and long-interval CoronaVac/BNT162b2 schedules found the long-interval regimen to have elicited higher anti-RBD immunoglobulin and IgG and higher neutralizing activities against alpha and beta SARS-CoV-2 variants than the short-interval regimen. These results align with other COVID-19 vaccine studies which showed increased antibody responses with an extended prime-boost interval [5, 6]. However, the long-interval heterologous regimen as a COVID-19 vaccination option has not been publicly recommended due to the fact that one dose immunisation by certain vaccines may not be effective in preventing the disease [7], thus leaving the one-dose vaccinated individuals at risk. Amidst the COVID-19 pandemic, the goal of vaccination was to ramp up the protective immunity among the population with a two-dose regimen in a short amount of time.

Aside from heightened immunoglobulin titters and inhibition percentages of neutralizing activities in BNT162b2-incorporated regimens compared to the CoronaVac/CoronaVac regimen, a notable discrepancy of anti-S1 IgA was observed. Serum anti-spike-1 protein-specific IgA OD/CO ratios were detected only in the BNT162b2-incorporated regimen. Limited data is available on the clinical benefit of serum IgA in protection; however, a recent in vitro experimental study has shown that serum IgA contributed to the neutralization of the SARS-CoV-2 [8-10].

Our study had a few noteworthy limitations. Firstly, the demographic discrepancies of the short and long-interval heterologous regimen cohorts heavily favour young females. Cohort sizes were relatively small, therefore requiring further studying with a larger sample size. Secondly, this study did not investigate the reactogenicity of the heterologous schedule. Lastly, efficacy data from more extensive trials are needed to comprehensively determine the benefits of heterologous CoronaVac followed by BNT162b2 regimen across all age groups in different countries facing different emerging SARS-CoV-2 variants.

## Conclusions

In low- and middle-income countries experiencing a vaccine shortage and emerging variants, heterologous COVID-19 vaccine schedules have the potential to accelerate vaccine rollout. Additionally, adverse events following vaccination called into question whether a different combination would be advantageous in reducing the chance of adverse reactions following the second dose of some vaccines, for example, mRNA vaccine which is related to myocarditis more frequently after the second than first dose vaccination [11]. Results of this preliminary investigation call for a larger safety and efficacy trial especially in males between 16-29 years of age, due to possible cases of myocarditis following the second dose of mRNA vaccine [12]. Further experimentation into T-cells responses is also of interest to fully elucidate immunological response to the heterologous regimen.

## Data Availability

All data produced in the present work are contained in the manuscript

## Acknowledgements

This work was supported by The National Research Council of Thailand (NRCT), Health Systems Research Institute, The Center of Excellence in Clinical Virology of Chulalongkorn University, and King Chulalongkorn Memorial Hospital. Nungruthai Suntronwong was supported by the second Century Fund (C2F) fellowship of Chulalongkorn University. We would like to thank Dr. Pussadee Hiranas, MD from the Navy Health Center. Somdechphrapinklao Hospital for the contribution to this project.

## Reference

[1] Wanlapakorn N, Suntronwong N, Phowatthanasathian H, et al. Safety and immunogenicity of heterologous and homologous inactivated and adenoviral-vectored COVID-19 vaccines in healthy adults. medRxiv 2021.11.04.21265908 [Preprint]. 2021 [cited 2021 Nov 6] Available from: https://www.medrxiv.org/content/10.1101/2021.11.04.21265908v2 https://doi.org/10.1101/2021.11.04.21265908

[2] Fadlyana E, Rusmil K, Tarigan R, et al. A phase III, observer-blind, randomized, placebo-controlled study of the efficacy, safety, and immunogenicity of SARS-CoV-2 inactivated vaccine in healthy adults aged 18-59 years: An interim analysis in Indonesia. Vaccine. 2021 Oct 22;39(44):6520–6528. doi: 10.1016/j.vaccine.2021.09.052. Epub 2021 Sep 24. PMID: 34620531; PMCID: PMC8461222.

[3] Tanriover MD, Doğanay HL, Akova M, et al. Efficacy and safety of an inactivated whole-virion SARS-CoV-2 vaccine (CoronaVac): interim results of a double-blind, randomised, placebo-controlled, phase 3 trial in Turkey. Lancet. 2021 Jul 17;398(10296):213–222. doi: 10.1016/S0140-6736(21)01429-X. Epub 2021 Jul 8. PMID: 34246358; PMCID: PMC8266301.

[4] Feng S, Phillips DJ, White T, et al. Correlates of protection against symptomatic and asymptomatic SARS-CoV-2 infection. Nat Med. 2021 Sep 29. doi: 10.1038/s41591-021-01540-1. Epub ahead of print. PMID: 34588689.

[5] Rogawski McQuade ET, Breskin A. Longer intervals and extra doses of ChAdOx1 nCoV-19 vaccine. Lancet. 2021 Sep 11;398(10304):933–935. doi: 10.1016/S0140-6736(21)01817-1. Epub 2021 Sep 1. PMID: 34480860; PMCID: PMC8409898.

[6] Zhang Y, Zeng G, Pan H, et al. Safety, tolerability, and immunogenicity of an inactivated SARS-CoV-2 vaccine in healthy adults aged 18-59 years: a randomised, double-blind, placebo-controlled, phase 1/2 clinical trial. Lancet Infect Dis. 2021 Feb;21(2):181–192. doi: 10.1016/S1473-3099(20)30843-4. Epub 2020 Nov 17. PMID: 33217362; PMCID: PMC7832443.

[7] Jara A, Undurraga EA, González C, et al. Effectiveness of an Inactivated SARS-CoV-2 Vaccine in Chile. N Engl J Med. 2021 Sep 2;385(10):875–884. doi: 10.1056/NEJMoa2107715. Epub 2021 Jul 7. PMID: 34233097; PMCID: PMC8279092.

[8] Sterlin D, Mathian A, Miyara M, et al. IgA dominates the early neutralizing antibody response to SARS-CoV-2. Sci Transl Med. 2021 Jan 20;13(577):eabd2223. doi: 10.1126/scitranslmed.abd2223. Epub 2020 Dec 7. PMID: 33288662; PMCID: PMC7857408.

[9] Klingler J, Weiss S, Itri V, et al. Role of Immunoglobulin M and A Antibodies in the Neutralization of Severe Acute Respiratory Syndrome Coronavirus 2. J Infect Dis. 2021 Mar 29;223(6):957–970. doi: 10.1093/infdis/jiaa784. PMID: 33367897; PMCID: PMC7798948.

[10] Wang Z, Lorenzi JCC, Muecksch F, et al. Enhanced SARS-CoV-2 neutralization by dimeric IgA. Sci Transl Med. 2021 Jan 20;13(577):eabf1555. doi: 10.1126/scitranslmed.abf1555. Epub 2020 Dec 7. PMID: 33288661; PMCID: PMC7857415.

[11] Gargano JW, Wallace M, Hadler SC, et al. Use of mRNA COVID-19 Vaccine After Reports of Myocarditis Among Vaccine Recipients: Update from the Advisory Committee on Immunization Practices - United States, June 2021. MMWR Morb Mortal Wkly Rep. 2021 Jul 9;70(27):977–982. doi: 10.15585/mmwr.mm7027e2. PMID: 34237049; PMCID: PMC8312754.

[12] Witberg G, Barda N, Hoss S, et al. Myocarditis after Covid-19 Vaccination in a Large Health Care Organization. N Engl J Med. 2021 Oct 6:NEJMoa2110737. doi: 10.1056/NEJMoa2110737. Epub ahead of print. PMID: 34614329; PMCID: PMC8531986.

